# Bivalent COVID-19 mRNA booster vaccination (BA.1 or BA.4/BA.5) increases neutralization of matched Omicron variants

**DOI:** 10.1101/2023.04.20.23288813

**Authors:** David N. Springer, Michael Bauer, Iris Medits, Jeremy V. Camp, Stephan W. Aberle, Clemens Burtscher, Eva Höltl, Lukas Weseslindtner, Karin Stiasny, Judith H. Aberle

**Affiliations:** Center for Virology, Medical University of Vienna, Austria; Health Center Erste Bank, Erste Bank, Vienna, Austria; Center for Public Health, Medical University of Vienna, Austria

**Keywords:** Bivalent COVID-19 booster vaccine, neutralizing antibodies, Omicron Breakthrough infection

## Abstract

We report SARS-CoV-2 neutralizing antibody titers in sera of triple-vaccinated individuals who received a booster dose of an original monovalent or a bivalent BA.1- or BA.4/BA.5-adapted vaccine, or had a breakthrough infection with Omicron variants BA.1, BA.2 or BA.4/BA.5. A bivalent BA.4/BA.5 booster or Omicron-breakthrough infection induced increased Omicron-neutralization titers compared with the monovalent booster. The XBB.1.5 variant effectively evaded neutralizing-antibody responses elicited by current vaccines and/or infection with previous variants.

## Text

The SARS-CoV-2 Omicron variant (B.1.1.529) evolved into several sublineages (BA.1 to BA.5, their descendants and recombinant forms), carrying mutations in the spike protein that result in escape from neutralizing antibodies elicited by ancestral vaccines^1^. Since late 2022, bivalent COVID-19 mRNA-booster vaccines, containing equal amounts of mRNAs encoding the ancestral (Wuhan-1) or Omicron spike proteins (BA.1 or BA.4/BA.5), are in widespread use. It is, however, still unclear whether a single booster dose with a bivalent vaccine would enhance neutralization of Omicron variants beyond that of original vaccines.

We investigated serum neutralization of SARS-CoV-2 variants after a bivalent BA.1 (n=12) or bivalent BA.4/BA.5 (n=22) booster of individuals who had previously received three vaccine doses. As reference cohorts, we included vaccinees with one (n=31) or two booster doses (n=26) of the original monovalent mRNA vaccine. Sera were collected 20-31d (bivalent-BA.1), 21-30d (bivalent-BA.4/BA.5), 15-43d (three-dose monovalent) and 16-38d (four-dose monovalent) after vaccination (Supplementary Tables 1 and 2). All specimens were nucleocapsid-antibody negative, indicating that these individuals had no previous SARS-CoV-2 infection. Serum-neutralizing activity was determined in a well-characterized live-virus neutralization test (NT)^2^, using SARS-CoV-2 wildtype (wt, D614G virus), Delta and Omicron variants (BA.1, BA.2, BA.5, XBB.1.5). We found for all cohorts that NT titers were highest against D614G (Figure 1a). The bivalent-BA.4/BA.5 booster induced significantly higher neutralization titers to BA.5 than the three- and four-dose monovalent boosters (Figure 1a). Consistent with our data, there is epidemiological evidence indicating that bivalent vaccines provide significant additional protection against symptomatic Omicron infection and hospitalization^3, 4^. Of note, our results showed only modestly increased BA.5 neutralization, which is in agreement with previous studies^5-12^, whereas others suggested that the bivalent-BA.4/BA.5 vaccine would not be more effective than the original monovalent vaccine^13-15^. The bivalent-BA.1 booster induced similarly high neutralization titers as the bivalent-BA.4/BA.5 booster (Figure 1a), but significant differences were not observed relative to the monovalent vaccine, which is in line with previous studies^6^. XBB.1.5-NT titers were lowest in all groups (Figure 1a), which confirms the strong escape of this variant from current vaccine- and infection-elicited neutralizing antibodies^8, 9, 16^

**Fig. 1.**
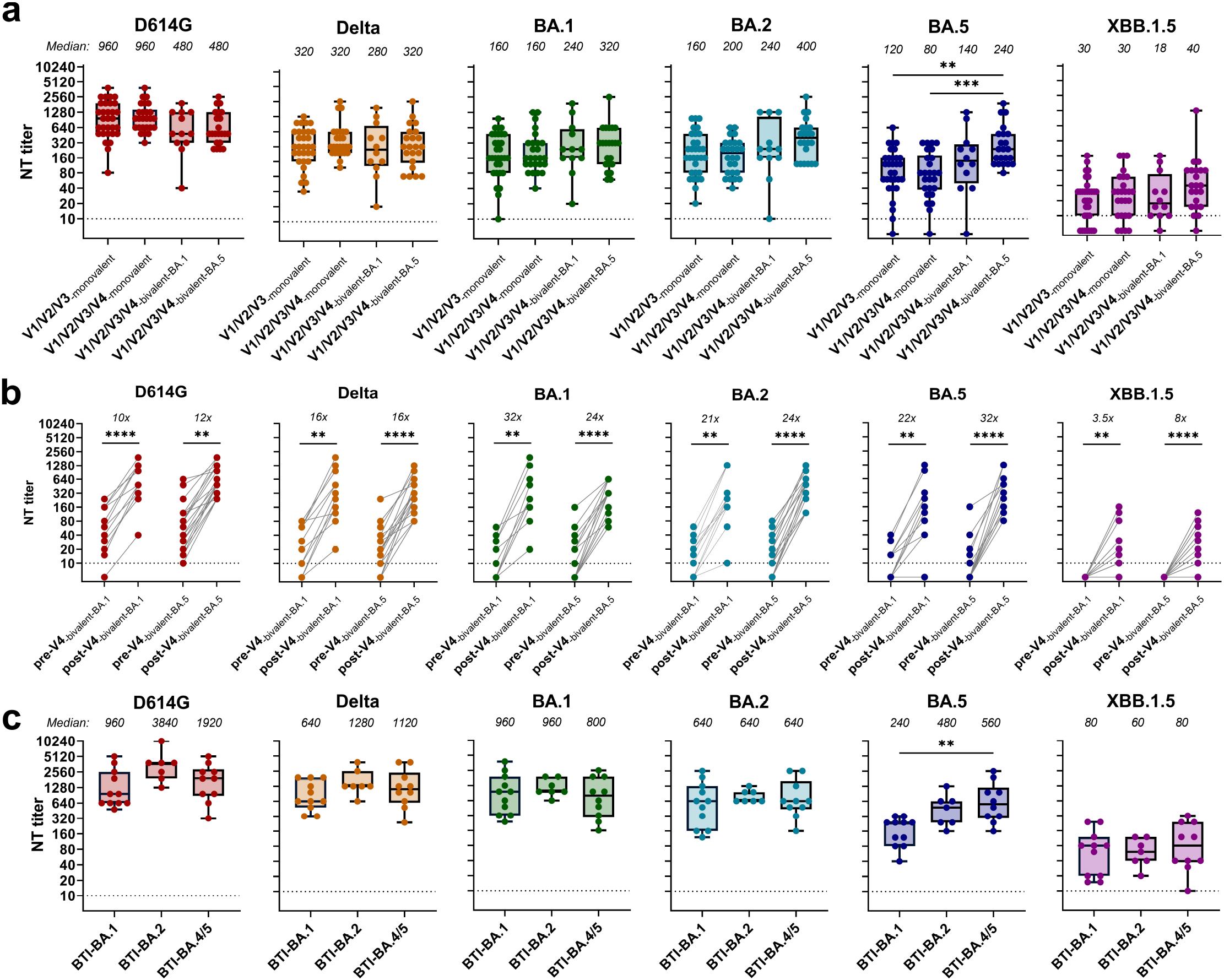
Serum-neutralizing activity against SARS-CoV-2 variants of vaccinated or breakthrough-infection cohorts. (a) NT titers of sera from individuals 3-4 weeks after a third or fourth dose of monovalent mRNA vaccines (V1/V2/V3-monovalent, V1/V2/V3/V4-monovalent), or bivalent-BA.1- or BA.4/BA.5-based mRNA vaccines (V1/V2/V3/V4-bivalent-BA.1, V1/V2/V3/V4- bivalent-BA.5). (b) NT titers at baseline (day 0, pre-V4) and after bivalent-BA.1 or BA.4/BA.5 booster vaccination (3-4 weeks, post-V4). (c) NT titers of sera from individuals with breakthrough infection with Omicron BA.1, BA.2 or BA.4/BA.5, following 2-4 doses of mRNA vaccines. Boxes range from 25th to 75th percentile, whiskers show min and max, and horizontal lines the median. BTI, breakthrough infection. NT titers were compared with Kruskal-Wallis test with Dunn’s multiple comparison correction. Paired data were analyzed with Wilcoxon’s signed-rank test followed by Bonferroni correction. ***p < 0.001, **p < 0.01.

A comparison of NT titers between sera obtained on the day of booster and those one month later showed that the bivalent-BA.1 vaccine yielded the strongest increase in BA.1-NT titers (Figure 1b). Similarly, the enhancement of NT titers of the bivalent-BA.4/BA.5 booster sera was greatest against BA.4/BA.5 (Figure 1b), indicating that variant-specific neutralizing antibodies were elicited by both bivalent vaccines.

We next analyzed sera from individuals after breakthrough infection with Omicron BA.1 (n=11), BA.2 (n=7) or BA.4/BA.5 (n=10) variants, following 2-4 doses of mRNA vaccination. Sera were obtained 13-36d, 16-25d and 22-52d after infection, respectively (Supplementary Table 3). NT titers after breakthrough infection were higher than after vaccination of SARS-CoV-2-naive individuals. BA.4/BA.5-breakthrough infections elicited significantly higher BA.5-NT titers than BA.1-breakthrough infections (Figure 1c). As for the vaccinees, XBB.1.5-NT titers were the lowest in all breakthrough cohorts (Figure 1c).

We used the NT titers (Figure 1) to generate antigenic maps that depict antigenic distances between all tested variants (Figure 2a). Variant distribution was similar for three- and four-dose-monovalent vaccine sera, with the highest reactivity around D614G, whereas the distance to BA.5 was approximately four antigenic units, equivalent to an 8-fold difference in neutralization (Figure 2). These data are in agreement with studies using post-vaccination sera with similar distributions of pre-Omicron and Omicron variants^11, 17^. The bivalent vaccine sera and Omicron-breakthrough infection sera yielded antigenic maps in which pre-Omicron and Omicron BA.1, BA.2 and BA.5 variants clustered tightly together, indicating increased neutralization breadth. Omicron XBB.1.5 mapped the furthest from the D614G strain, and the distance to XBB.1.5 was highest for monovalent booster sera (up to 5.5 antigenic units), followed by bivalent-BA.1 booster sera, and lowest for the bivalent-BA.5 booster sera (4 antigenic units). Additionally, we analyzed cumulative antigenic distance scores calculated from the sum of antigenic units between D614G and each variant measured for each serum (Figure 2b). In comparison to the monovalent vaccine sera, and consistent with the antigenic maps, these scores were significantly lower for bivalent-BA.1 and BA.4/BA.5 vaccine and breakthrough post-infection sera (Figure 2b). These data indicate that neutralizing activities were broader after bivalent BA.1 or BA.4/BA.5 boosters than after monovalent boosters.

**Fig. 2.**
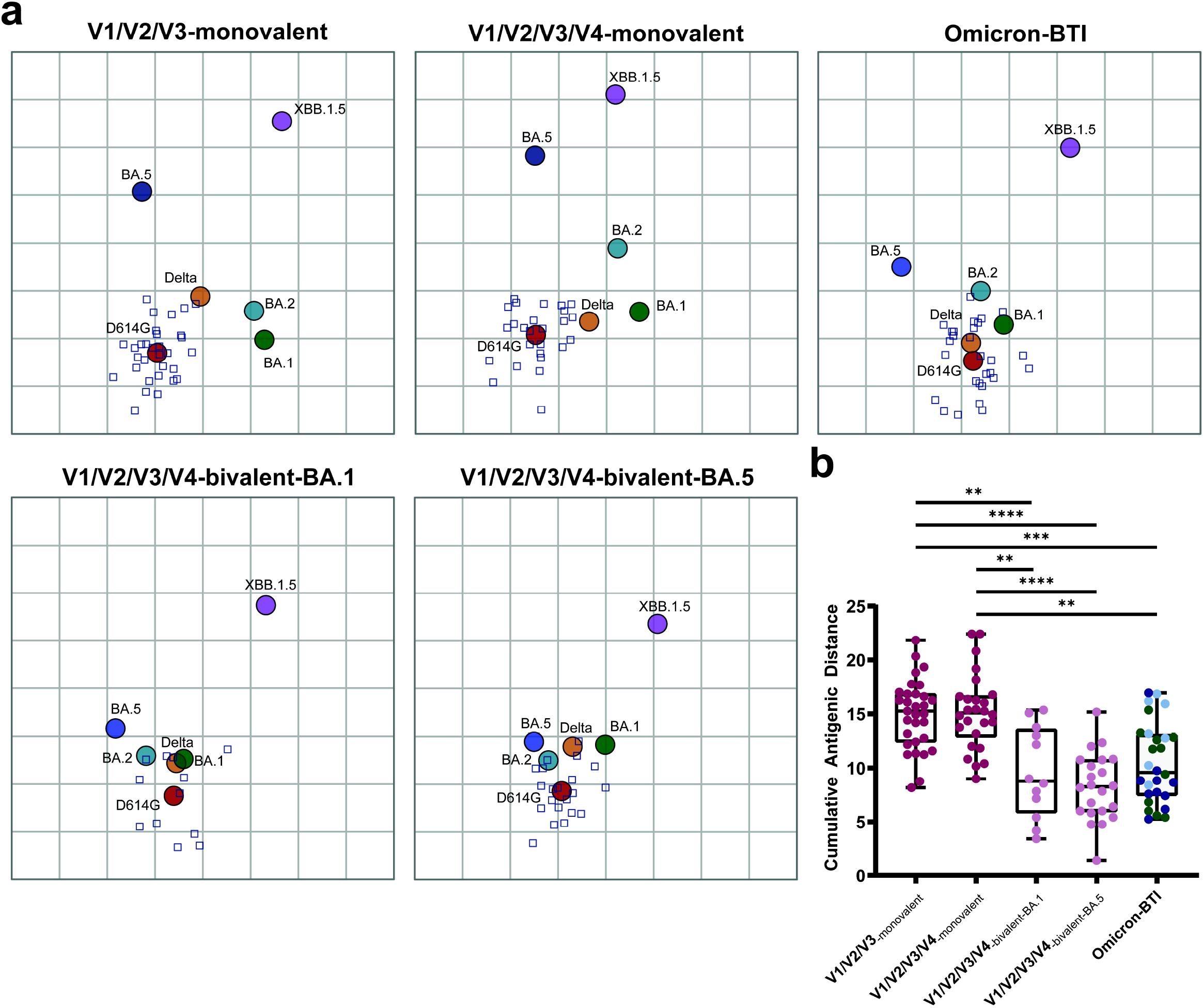
Antigenic maps and cumulative distance scores of SARS-CoV-2 variants for vaccinated and breakthrough-infection cohorts. (a) Antigenic maps of SARS-CoV-2 variants based on post-vaccination (V1/V2/V3-monovalent, n=31; V1/V2/V3/V4-monovalent, n=26; V1/V2/V3/V4-bivalent-BA.1, n=12; V1/V2/V3/V4-bivalent-BA.5, n=22) and Omicron breakthrough-infection sera, including subvariants BA.1, BA.2 and BA.4/BA.5 (n=22). Squares represent individual sera, circles SARS-CoV-2 variants. The x- and y-axes of the maps are antigenic distances, and each square represents a two-fold change in neutralization titer. (b) Cumulative antigenic distance scores. Boxes range from 25th to 75th percentile, whiskers show min and max, and horizontal lines the median. BTI, breakthrough infection. Scores were compared with Kruskal-Wallis tests with Dunn’s multiple comparison correction. ****p < 0.0001, ***p < 0.001.

Our study has several limitations: First, the sample size was relatively small with some differences among study groups (Supplementary Tables 1-3), including (i) unequal numbers of vaccine doses, i.e., more participants with 4 doses among the BA.4/BA.5-breakthrough-infection cohort; (ii) some participants in the bivalent but not in the monovalent cohort had received vector vaccines as a first dose; (iii) more female than male participants in monovalent (3-dose) vs. bivalent vaccine groups. Second, we did not measure non-neutralizing antibodies and cellular responses that are likely involved in durable protection against severe COVID-19, with T cell responses being highly cross-reactive against Omicron and prior variants^1^. Third, we do not know how NT titers relate to protection against infection, severe disease and death^1, 18^

In summary, our data show that a bivalent booster elicits broader Omicron-neutralizing activities and are concordant with recent real-world data demonstrating superior protection against severe disease by BA.5-bivalent booster vaccines^3^. Moreover, our data support the conclusion that the recently emerged XBB.1.5 variant effectively evades neutralizing-antibody responses elicited by current vaccines or breakthrough infection with previously circulating variants.

## METHODS

### Omicron variant identification

Nasopharyngeal swabs were analyzed with the mutation assays VirSNiP SARS-CoV-2 Spike S371L S373P, VirSNiP SARS-CoV-2 Spike 484A 486V and VirSNiP SARS-CoV-2 Spike L452R (TIB MOLBIOL, Berlin, Germany). Characteristic melting peaks for the mutations S371L & S373P indicated an infection with Omicron BA.1, S371F & S373P indicated BA.2, and S371F & S373P with the additional mutations L452R & F486V indicated BA.4/5, respectively.

### SARS-CoV-2 neutralization test (NT)

SARS-CoV-2 strains were isolated from nasopharyngeal swabs of infected individuals using Vero E6 (ECACC #85020206) or Vero E6-TMPRSS2 cells (kindly provided by Anna Ohradanova-Repic), as described previously^2^. Sequences determined by next-generation sequencing were uploaded to the GSAID database (wt, B.1.1 with the D614G mutation:EPI_ISL_438123; Delta, B.1.617.2-like, sub-lineage AY.122:EPI_ISL_4172121; Omicron, B.1.1.529+BA.*, sub-lineage BA.1.17:EPI_ISL_9110894; Omicron, B.1.1.529+BA.*, sublineage BA.2:EPI_ISL_11110193; Omicron, B.1.1.529+BA.*, sub-lineage BA.5.3:EPI_ISL_15982848; XBB.1.5:EPI_ISL_17062381. Pango lineages were determined with Pango v.4.1.3, Pango-data v1.17.).

Live-virus NTs were performed as previously described^2^. Serial two-fold dilutions of heat-inactivated serum (duplicates) were incubated with 50-100 TCID_50_ SARS-CoV-2 for 1h at 37°C. The sample-virus mixtures were added to Vero E6 cells and incubated for three to five days at 37°C. NT titers were expressed as the highest reciprocal serum dilution that prevented cytopathic effect, which was assessed microscopically. NT titers ≥10 were considered positive.

### Antigenic cartography

We constructed antigenic maps based on serum-neutralization data with antigenic cartography^11, 17^. The position of the variants and sera corresponds to the fold-difference to the maximum titer for each serum. A grid unit in any direction (one antigenic unit) represents a two-fold change in neutralization titer. Antigenic maps were generated with the Racmacs package (https://github.com/acorg/Racmacs)^19^ in R with 500 optimization steps and the minimum column basis parameter set to ‘none’.

### Ethics

All work was conducted in accordance with the Declaration of Helsinki in terms of informed consent and approval by an appropriate institutional board. Written informed consent from study participants was not required, as the analysis was performed on anonymized leftover samples from routine laboratory diagnosis in accordance with national legislation and institutional requirements. The ethics committee of the Medical University of Vienna, Austria, approved the study protocol (EK1035/2016, EK1513/2016, EK1926/2020, EK1291/2021).

### Statistical analyses

Statistical analysis was performed with Graphpad Prism 9.3.1 and R 4.2.0. Kruskal-Wallis test (two-tailed) with Dunn’s multiple comparison correction was used to compare NT titers and cumulative antigenic distance scores between different cohorts. For medians and fold-changes of pre- and post-vaccination titers, values <10 were set to 5. Wilcoxon’s signed-rank test followed by Bonferroni correction was used to compare paired data. Alpha was set to 0.05.

## Data Availability

All data produced in the present study are available upon reasonable request to the authors

## DATA AVAILABILITY

The datasets generated during and/or analyzed during the current study are available from the corresponding authors on reasonable request.

## ACKNOWLEDGEMENTS

We thank Jutta Hutecek, Sylvia Malik, Barbara Dalmatiner, Katja Prüger and Elke Peil for their excellent technical assistance.

## AUTHOR CONTRIBUTIONS

JA, KS: conceptualization. JA, KS, DS: writing of the manuscript. KS, JA, JC, SA, LW, DS, MB, IM: methodology and experimental work. JA, KS, LW, EH, CB: resources. DS, MB, KS: data analysis.

JA: funding acquisition. KS and JA: supervision. All authors contributed to the manuscript and approved the submitted version.

## COMPETING INTEREST

The authors have no competing interests to declare.

## Notes

### Competing Interest Statement

The authors have declared no competing interest.

### Funding Statement

This study did not receive any funding

### Author Declarations

The ethics committee of the Medical University of Vienna, Austria, gave ethical approval for this work.

## REFERENCES

1. Barouch, D.H. Covid-19 Vaccines - Immunity, Variants, Boosters. N Engl J Med 387, 1011–1020 (2022).

2. Medits, I. et al. Different Neutralization Profiles After Primary SARS-CoV-2 Omicron BA.1 and BA.2 Infections. Front Immunol 13, Article 946318 (2022).

3. Lin, D.Y. et al. Effectiveness of Bivalent Boosters against Severe Omicron Infection. N Engl J Med 388, 764–766 (2023).

4. Link-Gelles R, C.A., Fleming-Dutra KE, et al. Effectiveness of Bivalent mRNA Vaccines in Preventing Symptomatic SARS-CoV-2 Infection — Increasing Community Access to Testing Program, United States, September–November 2022. MMWR Morb Mortal Wkly Rep 2022;71:1526–1530 (2022).

5. Addetia, A. et al. Therapeutic and vaccine-induced cross-reactive antibodies with effector function against emerging Omicron variants. bioRxiv (2023).

6. Chalkias, S. et al. A Bivalent Omicron-Containing Booster Vaccine against Covid-19. N Engl J Med 387, 1279–1291 (2022).

7. Davis-Gardner, M.E. et al. Neutralization against BA.2.75.2, BQ.1.1, and XBB from mRNA Bivalent Booster. N Engl J Med (2022).

8. Jiang, N. et al. Bivalent mRNA vaccine improves antibody-mediated neutralization of many SARS-CoV-2 Omicron lineage variants. bioRxiv, 2023.2001.2008.523127 (2023).

9. Kurhade, C. et al. Low neutralization of SARS-CoV-2 Omicron BA.2.75.2, BQ.1.1 and XBB.1 by parental mRNA vaccine or a BA.5 bivalent booster. Nat Med 29, 344–347 (2023).

10. Qu, P. et al. Extraordinary Evasion of Neutralizing Antibody Response by Omicron XBB.1.5, CH.1.1 and CA.3.1 Variants. bioRxiv, 2023.2001.2016.524244 (2023).

11. Wang, W. et al. Antigenic cartography of well-characterized human sera shows SARS-CoV-2 neutralization differences based on infection and vaccination history. Cell Host Microbe 30, 1745–1758 e1747 (2022).

12. Zou, J. et al. Improved Neutralization of Omicron BA.4/5, BA.4.6, BA.2.75.2, BQ.1.1, and XBB.1 with Bivalent BA.4/5 Vaccine. bioRxiv, 2022.2011.2017.516898 (2022).

13. Hoffmann, M. et al. Effect of hybrid immunity and bivalent booster vaccination on omicron sublineage neutralisation. Lancet Infect Dis 23, 25–28 (2023).

14. Wang, Q. et al. Antibody Response to Omicron BA.4-BA.5 Bivalent Booster. New Engl J Med (2023).

15. Collier, A.Y. et al. Immunogenicity of BA.5 Bivalent mRNA Vaccine Boosters. N Engl J Med 388, 565–567 (2023).

16. Hoffmann, M. et al. Profound neutralization evasion and augmented host cell entry are hallmarks of the fast-spreading SARS-CoV-2 lineage XBB.1.5. Cell Mol Immunol, 1–4 (2023).

17. van der Straten, K. et al. Antigenic cartography using sera from sequence-confirmed SARS-CoV-2 variants of concern infections reveals antigenic divergence of Omicron. Immunity 55, 1725–1731 e1724 (2022).

18. Krammer, F. A correlate of protection for SARS-CoV-2 vaccines is urgently needed. Nat Med 27, 1147–1148 (2021).

19. Wilks, S.H. et al. Mapping SARS-CoV-2 antigenic relationships and serological responses. bioRxiv, 2022.2001.2028.477987 (2022).

