## Supplemental Table 1-3 for "Bivalent COVID-19 mRNA booster vaccination (BA.1 or BA.4/BA.5) increases neutralization of matched Omicron variants"

### Supplementary Information:

**Supplementary Table 1: Vaccination history of the monovalent vaccinated individuals**

| *Study-ID (Person)* | *Age (years)* | *Sex* | *1^st^  vacc.* | *2^nd^  vacc.* | *3^rd^  vacc.* | *4^th^  vacc.* | *Days 2^nd^ vacc to 3^rd^ vacc.* | *Days 3^rd^ vacc to 4^th^ vacc.* | *Days 3^rd^ vacc. to blood sampling* | *Days 4^th^ vacc. to blood sampling* |
| --- | --- | --- | --- | --- | --- | --- | --- | --- | --- | --- |
| *P3* | *50-54* | *f* | *P* | *P* | *P* | *N* | 285 | *N* | *26* | *N* |
| *P4* | *25-29* | *f* | *P* | *P* | *P* | *N* | 288 | *N* | *23* | *N* |
| *P5* | *45-49* | *f* | *P* | *P* | *P* | *N* | 287 | *N* | *20* | *N* |
| *P9* | *50-54* | *f* | *P* | *P* | *P* | *N* | 287 | *N* | *29* | *N* |
| *P13* | *30-34* | *f* | *P* | *P* | *P* | *N* | 290 | *N* | *26* | *N* |
| *P14* | *50-54* | *m* | *P* | *P* | *P* | *N* | 273 | *N* | *24* | *N* |
| *P15* | *45-49* | *f* | *P* | *P* | *P* | *N* | 287 | *N* | *24* | *N* |
| *P36* | *25-29* | *f* | *P* | *P* | *P* | *N* | 244 | *N* | *38* | *N* |
| *P17* | *45-49* | *f* | *P* | *P* | *M* | *N* | 224 | *N* | *38* | *N* |
| *P19* | *30-34* | *f* | *P* | *P* | *P* | *N* | 301 | *N* | *43* | *N* |
| *P22* | *30-34* | *f* | *P* | *P* | *M* | *N* | 224 | *N* | *28* | *N* |
| *P23* | *35-39* | *f* | *P* | *P* | *M* | *N* | 211 | *N* | *33* | *N* |
| *P24* | *35-39* | *f* | *P* | *P* | *M* | *N* | 224 | *N* | *28* | *N* |
| *P25* | *55-59* | *f* | *P* | *P* | *P* | *N* | 245 | *N* | *22* | *N* |
| *P26* | *25-29* | *f* | *P* | *P* | *P* | *N* | 268 | *N* | *21* | *N* |
| *P37* | *25-29* | *m* | *P* | *P* | *P* | *N* | 192 | *N* | *26* | *N* |
| *P35* | *25-29* | *m* | *P* | *P* | *P* | *N* | 278 | *N* | *23* | *N* |
| *P1* | *50-54* | *f* | *P* | *P* | *P* | *P* | 246 | 265 | *N* | *20* |
| *P2* | *40-44* | *m* | *P* | *P* | *P* | *P* | 287 | 245 | *N* | *16* |
| *P42* | *35-39* | *f* | *P* | *P* | *M* | *P* | 238 | 274 | *N* | *20* |
| *P6* | *50-54* | *f* | *P* | *P* | *M* | *P* | 235 | 287 | *28* | *20* |
| *P8* | *55-59* | *f* | *P* | *P* | *P* | *P* | 287 | 240 | *28* | *25* |
| *P10* | *35-39* | *f* | *P* | *P* | *P* | *P* | 286 | 265 | *21* | *23* |
| *P11* | *50-54* | *f* | *P* | *P* | *M* | *P* | 212 | 152 | *32* | *31* |
| *P12* | *55-59* | *f* | *P* | *P* | *P* | *M* | 163 | 175 | *40* | *29* |
| *P16* | *25-29* | *f* | *P* | *P* | *P* | *P* | 287 | 224 | *15* | *21* |
| *P18* | *50-54* | *f* | *P* | *P* | *M* | *P* | 238 | 300 | *22* | *30* |
| *P21* | *45-49* | *m* | *P* | *P* | *P* | *P* | 288 | 229 | *16* | *21* |
| *P27* | *60-64* | *f* | *P* | *P* | *P* | *P* | 287 | 156 | *30* | *22* |
| *P28* | *80-84* | *f* | *P* | *P* | *P* | *P* | 205 | 147 | *N* | *38* |
| *P43* | *80-84* | *m* | *M* | *M* | *M* | *M* | NA | 211 | *N* | *26* |
| *P29* | *55-59* | *m* | *P* | *P* | *P* | *P* | 245 | 262 | *N* | *24* |
| *P38* | *40-44* | *f* | *P* | *P* | *P* | *P* | NA | 231 | *N* | *21* |
| *P39* | *35-39* | *m* | *P* | *P* | *P* | *P* | 75 | 267 | *N* | *24* |
| *P30* | *50-54* | *f* | *P* | *P* | *P* | *M* | 251 | 251 | *30* | *30* |
| *P44* | *55-59* | *m* | *P* | *P* | *P* | *P* | 185 | 221 | *N* | *24* |
| *P45* | *55-59* | *f* | *P* | *P* | *P* | *P* | 128 | 228 | *N* | *22* |
| *P46* | *65-69* | *m* | *P* | *P* | *P* | *P* | 185 | 262 | *N* | *22* |
| *P31* | *55-59* | *f* | *P* | *P* | *P* | *M* | 252 | 263 | *32* | *24* |
| *P32* | *50-54* | *f* | *P* | *P* | *P* | *P* | 226 | 292 | *32* | *25* |
| *P33* | *60-64* | *m* | *P* | *P* | *P* | *P* | 283 | 241 | *21* | *21* |
| *P34* | *60-64* | *f* | *P* | *P* | *P* | *P* | 260 | 270 | *27* | *29* |
| *P41* | *55-59* | *f* | *P* | *P* | *P* | *P* | 180 | 274 | *N* | *18* |

*Age: in years at time of blood sampling; m: male. f: female; vacc.: vaccination; P: Biontech/Pfizer “Comirnaty” BNT162b2; M: Moderna “Spikevax” mRNA-1273; NA: not available, N: No*

*Study-IDs do not reveal the identity of the study subjects.*

**Supplementary Table 2: Vaccination history of the bivalent vaccinated individuals**

| *Study-ID (Person)* | *Age (years)* | *Sex* | *1^st^ vacc.* | *2^nd^ vacc.* | *3^rd^ vacc.* | *4^th^ vacc.* | *Days 2^nd^ vacc to 3^rd^ vacc.* | *Days 3^rd^ vacc. to 4^th^ vacc.* | *Days 4^th^ vacc. to blood sampling* |
| --- | --- | --- | --- | --- | --- | --- | --- | --- | --- |
| *P89* | *35-39* | *m* | *AZ* | *AZ* | *P* | *P-BA.1* | 177 | 306 | 27 |
| *P90* | *50-54* | *f* | *P* | *P* | *P* | *P-BA.1* | 142 | 306 | 21 |
| *P91* | *50-54* | *f* | *P* | *P* | *P* | *P-BA.1* | 160 | 308 | 28 |
| *P92* | *35-39* | *m* | *P* | *P* | *P* | *P-BA.1* | 146 | 291 | 21 |
| *P93* | *60-64* | *m* | *P* | *P* | *P* | *P-BA.1* | 162 | 291 | 21 |
| *P94* | *50-54* | *m* | *P* | *P* | *P* | *P-BA.1* | *NA* | *NA* | 21 |
| *P95* | *50-54* | *f* | *AZ* | *AZ* | *P* | *P-BA.1* | 138 | 302 | 31 |
| *P96* | *40-44* | w | J | P | P | *P-BA.1* | 148 | 203 | 21 |
| *P97* | *40-44* | m | P | P | P | *P-BA.1* | 148 | 279 | 22 |
| *P98* | *55-59* | m | P | P | P | *P-BA.1* | 167 | 308 | 21 |
| *P99* | *50-54* | m | P | P | P | *P-BA.1* | 277 | 238 | 21 |
| *P100* | *45-49* | w | P | P | P | *P-BA.1* | 138 | 296 | 20 |
| *P101* | *35-39* | *f* | *P* | *P* | *P* | *P-BA.5* | 147 | 331 | 21 |
| *P102* | *45-49* | *m* | *P* | *P* | *P* | *P-BA.5* | 157 | 309 | 21 |
| *P103* | *35-39* | *m* | *J* | *P* | *P* | *P-BA.5* | 125 | 231 | 21 |
| *P104* | *30-34* | *m* | *P* | *P* | *P* | *P-BA.5* | 107 | 243 | 21 |
| *P105* | *55-59* | *f* | *J* | *P* | *P* | *P-BA.5* | 122 | 232 | 21 |
| *P106* | *55-59* | *f* | *P* | *P* | *P* | *P-BA.5* | 187 | 302 | 21 |
| *P107* | *50-54* | *f* | *AZ* | *AZ* | *P* | *P-BA.5* | 138 | 308 | 21 |
| *P108* | *60-64* | *m* | *P* | *P* | *P* | *P-BA.5* | 183 | 298 | 21 |
| *P109* | *50-54* | *f* | *P* | *P* | *P* | *P-BA.5* | 188 | 306 | 21 |
| *P110* | *40-44* | *m* | *J* | *P* | *P* | *P-BA.5* | 125 | 232 | 21 |
| *P111* | *35-39* | *f* | *P* | *P* | *P* | *P-BA.5* | 134 | 301 | 21 |
| *P112* | *30-34* | *f* | *P* | *P* | *P* | *P-BA.5* | 132 | 318 | 21 |
| *P113* | *55-59* | *f* | *P* | *P* | *P* | *P-BA.5* | 140 | 298 | 21 |
| *P114* | *45-49* | *f* | *P* | *P* | *P* | *P-BA.5* | 173 | 287 | 21 |
| *P115* | *55-59* | *f* | *P* | *P* | *P* | *P-BA.5* | 158 | 304 | 21 |
| P116 | *50-54* | f | P | P | P | *P-BA.5* | 163 | 307 | 21 |
| P117 | *55-59* | f | P | P | P | *P-BA.5* | 148 | 298 | 21 |
| P118 | *50-54* | f | AZ | AZ | P | *P-BA.5* | 153 | 306 | 21 |
| P119 | *55-59* | m | J | P | P | *P-BA.5* | 105 | 243 | 21 |
| P120 | *55-59* | m | P | P | P | *P-BA.5* | 179 | 322 | 30 |
| BT55 | *25-29* | m | P | P | P | *P-BA.5* | 278 | 329 | 28 |
| P121 | *25-29* | f | P | P | P | *P-BA.5* | *NA* | 330 | 28 |

*Age: in years at time of blood sampling; m: male, f: female; vacc.: vaccination; AZ: Astra-Zeneca “Vaxzevria” ChAdOx1; P: Monovalent Biontech/Pfizer “Comirnaty” BNT162b2; J: Johnson & Johnson Ad26.COV2.S;* *P-BA.1: Biontech/Pfizer Bivalent (WT/BA.1) BNT162b2 BA.1; P-BA.5: Biontech/Pfizer Bivalent (WT/BA.5) BNT162b2 BA.5; M: Moderna “Spikevax” mRNA-1273; NA: not available. Study-IDs do not reveal the identity of the study subjects.*

**Supplementary Table 3: Vaccination history of the subjects who experienced Omicron breakthrough infection**

| *Study-ID (Person)* | *Age (years)* | *Sex* | *1^st^ vacc.* | *2^nd^ vacc.* | *3^rd^ vacc.* | *4^th^ vacc.* | *Days 2^nd^ vacc to 3^rd^ vacc.* | *Days 3rd vacc. to 4^th^ vacc.* | *Infecting Omicron variant* | *Days last vacc. to infection* | *Days infection to blood sampling* |
| --- | --- | --- | --- | --- | --- | --- | --- | --- | --- | --- | --- |
| *P09* | *50-54* | *f* | *P* | *P* | *P* | *N* | 287 | *N* | *BA.1* | *101* | *20* |
| *P17* | *45-49* | *f* | *P* | *P* | *M* | *N* | 224 | *N* | *BA.1* | *143* | *19* |
| *P22* | *30-34* | *f* | *P* | *P* | *M* | *N* | 224 | *N* | *BA.1* | *139* | *18* |
| *P23* | *35-39* | *f* | *P* | *P* | *M* | *N* | 211 | *N* | *BA.1* | *142* | *18* |
| *P52* | *25-29* | *m* | *M* | *M* | *M* | *N* | 185 | *N* | *BA.1* | *28* | *20* |
| *P53* | *20-24* | *m* | *P* | *P* | *P* | *N* | 178 | *N* | *BA.1* | *35* | *20* |
| *P54* | *25-29* | *f* | *P* | *P* | *N* | *N* | *N* | *N* | *BA.1* | *202* | *17* |
| *P55* | *40-44* | *m* | *P* | *P* | *N* | *N* | *N* | *N* | *BA.1* | *123* | *13* |
| *P56* | *75-79* | *f* | *P* | *P* | *P* | *N* | 195 | *N* | *BA.1* | *110* | *30* |
| *P57* | *30-34* | *f* | *P* | *P* | *P* | *N* | 145 | *N* | *BA.1* | *105* | *36* |
| *P59* | *55-59* | *m* | *P* | *P* | *P* | *N* | 165 | *N* | *BA.1* | *99* | *24* |
| *P04* | *25-29* | *f* | *P* | *P* | *P* | *N* | 288 | *N* | *BA.2* | *110* | *22* |
| *P28* | *80-84* | *f* | *P* | *P* | *P* | *P* | 205 | 147 | *BA.2* | *80* | *16* |
| *P37* | *25-29* | *m* | *P* | *P* | *P* | *N* | 192 | *N* | *BA.2* | *59* | *28* |
| *P48* | *35-39* | *f* | *P* | *P* | *M* | *N* | 238 | *N* | *BA.2* | *245* | *56* |
| *P49* | *55-59* | *f* | *P* | *P* | *M* | *N* | 224 | *N* | *BA.2* | *190* | *27* |
| *P51* | *55-59* | *f* | *P* | *P* | *P* | *N* | 244 | *N* | *BA.2* | *179* | *35* |
| *P58* | *50-54* | *f* | *P* | *P* | *P* | *N* | 143 | *N* | *BA.2* | *114* | *27* |
| *P03* | *50-54* | *f* | *P* | *P* | *P* | *N* | 285 | *N* | *BA.4/5* | *218* | *24* |
| *P14* | *50-54* | *m* | *P* | *P* | *P* | *N* | 273 | *N* | *BA.4/5* | *216* | *28* |
| *P19* | *30-34* | *f* | *P* | *P* | *P* | *N* | 301 | *N* | *BA.4/5* | *199* | *24* |
| *P24* | *35-39* | *f* | *P* | *P* | *M* | *N* | 224 | *N* | *BA.4/5* | *321* | *30* |
| *P25* | *60-64* | *f* | *P* | *P* | *P* | *P* | 245 | 282 | *BA.4/5* | *13* | *22* |
| *P27* | *60-64* | *f* | *P* | *P* | *P* | *P* | 287 | 156 | *BA.4/5* | *40* | *36* |
| *P29* | *55-59* | *m* | *P* | *P* | *P* | *P* | 245 | 262 | *BA.4/5* | *50* | *23* |
| *P44* | *55-59* | *m* | *P* | *P* | *P* | *P* | 185 | 221 | *BA.4/5* | *38* | *24* |
| *P47* | *45-49* | *f* | *P* | *P* | *M* | *N* | 218 | *N* | *BA.4/5* | *285* | *52* |
| *P50* | *55-59* | *f* | *P* | *P* | *P* | *N* | 288 | *N* | *BA.4/5* | *212* | *22* |

*Age: in years at time of blood sampling; m: male, f: female; vacc.: vaccination; AZ: Astra-Zeneca “Vaxzevria” ChAdOx1; P: Monovalent Biontech/Pfizer “Comirnaty” BNT162b2; P-BA.1: Biontech/Pfizer Bivalent (WT/BA.1) BNT162b2 BA.1; P-BA.5: Biontech/Pfizer Bivalent (WT/BA.5) BNT162b2 BA.5, M: Moderna “Spikevax” mRNA-1273; NA: not available, N: No. Study-IDs do not reveal the identity of the study subjects.*
